# Bioinformatical enrichment analysis of genes involved in the pathway of endometriosis disease

**DOI:** 10.1101/2024.02.09.24302561

**Authors:** Kusum Kusum, Ashish Ashish, Ravi Bhushan, Radha Chaube, Sangeeta Rai, Royana Singh

**Affiliations:** Department of Zoology, Institute of Science, Banaras Hindu University, Varanasi-221005 Uttar Pradesh, India; MRU Lab, Department of Anatomy, Institute of Medical Science, Banaras Hindu University, Varanasi, 221005, Uttar Pradesh, India; Department of Chromosomal Genetics Disorder Institute of Science, Banaras Hindu University, Varanasi-221005 Pradesh, India; Obstetrics and gynecology, Institute of Medical Science, Banaras Hindu University Varanasi-221005 Pradesh, India

**Keywords:** Endometriosis, mRNA seq analysis, reproductive women, estrogen, aromatase

## Abstract

Endometriosis is a gynecological disease characterized by the presence of uterine (eutopic) endometrial glands and tissues outside the intra-uterine locations, in ectopic regions such as the pelvic peritoneum, fallopian tubes, or ovaries. Approximately 5-10% of reproductive and 20-50% of infertile women are affected by endometriosis. The pathogenesis of endometriosis involves various factors, including hormonal, environmental, genetic, and immune system components, directly or indirectly altering estrogen levels and impacting women’s reproductive health.This study aimed to identify novel and potential biomarkers for endometriosis using mRNA seq analysis. Differentially expressed genes (DEGs) were identified from raw gene expression profiles, and their functional analysis was subsequently conducted. A total of 552 DEGs (312 upregulated and 240 downregulated) were identified in samples from women with endometriosis compared to control subjects.Major DEGs, such as C3, PSAP, APP, GNG12, were identified as hub nodes and found to be involved in various functions, including epithelial cell differentiation and development, proteolysis, gland development, muscle fiber development, and response to hormone stimulus. These DEGs may play a direct or indirect role in the pathogenesis of endometriosis, serving as potential biomarkers for ectopic endometrium. While this study provides a preliminary insight into the mechanism of endometriosis, further detailed studies are necessary to fully understand its path of action.

## Introduction

Endometriosis is a pronounced gynecological disease that significantly impacts women’s health. This disorder is characterized by the presence of endometrial glands and stromal tissues outside the uterine endometrium (eutopic region), extending to ectopic regions such as the pelvic peritoneum, fallopian tubes, or ovaries (1–3). Being an estrogen-dependent disorder, it affects approximately 5-10% of reproductively active women and 20-50% of women diagnosed with infertility globally (4).Despite its prevalence, the etiology and pathogenesis of endometriosis remain unclear, imposing economic and reproductive health burdens on affected women (5). Various theories have been proposed to explain its occurrence, with Sampson’s theory of retrograde menstruation suggesting the transportation of endometrial cells from eutopic regions to ectopic regions, leading to endometriosis (6). However, extensive studies are needed to distinguish expression patterns in eutopic and ectopic endometrium (7).The absence of a non-invasive diagnostic marker using serum, urine, or endometrial tissue samples highlights the urgent need for early disease detection (8). The precise mechanisms affecting the overall pathology of endometriosis remain unknown. The recent development of RNA-Seq analysis, a deep-sequencing technology, provides a novel approach to studying transcriptomes (9). Transcriptomes represent the complete set of transcripts in a cell under a specific physiological condition, offering insights into the functional elements at play (10).Molecular analysis comparing the endometrium of women with endometriosis to normal endometrium as a control holds promise for understanding the disease’s pathophysiology and identifying specific biomarkers for diagnosis.

## Materials and Methods

### Microarray Data and Samples

Raw gene expression profiles were retrieved from the National Center of Biotechnology Information (NCBI) Gene Expression Omnibus (GEO) database (http://www.ncbi.nlm.nih.gov/geo/) under the dataset ID GSE7305 (11). The dataset comprised samples from normal eutopic endometrium, as well as ectopic and eutopic endometrium from patients with endometriosis.

### Study Design

The study focused on analyzing genes associated with both eutopic and ectopic endometrium in samples from control subjects and patients with endometriosis. Bioinformatic tools were employed for a comprehensive analysis of the microarray data.

### Ethical Approval

This microarray study received approval from the ethical committee of the institution, Banaras Hindu University, under the reference number Dean/2018/EC/936.

### Data Processing and DEG Screening

In processing raw gene expression datasets, probe-specific expression values were averaged to derive gene expression values. The BiGGEsTs software analysis tool was then utilized to identify up and downregulated genes. Subsequently, GEO2R (https://www.ncbi.nlm.nih.gov/geo/geo2r/) was employed to convert probe-level symbols into gene-level symbols. All differentially expressed genes (DEGs) with p-values < 0.05 and threshold logFC values > 0.1 for upregulated genes and < −0.1 for downregulated genes were selected (12).

### Principal Component Analysis and Heat Map Generation

Principal component analysis (PCA) was conducted using the online tool ClustVis (13), specific lly for DEGs. Due to size limitations of ClustVis (supporting file sizes up to 2MB), a PCA plot for the total gene expression was not feasible.

### Identification of Novel Endometriosis Biomarkers

To identify novel biomarkers associated with endometriosis, the list of differentially expressed genes was compared against reported gene lists obtained from OMIM (https://www.omim.org/) and Gene Cards (https://www.genecards.org/) (14). Venny 2.1 (https://bioinfogp.cnb.csic.es/tools/venny/) was utilized for comparison and construction of Venn diagrams (15).

### Construction of Protein-Protein Interaction (PPI) Network and Sub-Network Mining

Differentially expressed genes (DEGs) were identified and uploaded to STRING v 10.5 (http://www.string-db.org/) (16), an online database predicting functional interactions between proteins. A combined score > 0.4 served as the baseline criteria for protein-protein interaction (PPI) among gene pairs. Subsequently, the network and sub-network were constructed using Cytoscape v3.2.1 (http://www.cytoscape.org/) (17), a software for visualizing and analyzing biological networks. The criteria for network construction included clustering coefficient and edge betweenness.

### Functional and Pathway Enrichment Analysis of DEGs

Gene ontology (GO) enrichment analysis, encompassing biological processes (BP), molecular functions (MF), and cellular components (CC), was performed using DAVID v 6.8 (database for annotation, visualization, and integrated discovery, http://david.abcc.ncifcrf.gov/) (18). This program integrates a comprehensive set of functional annotations for a large gene list. Based on hypergeometric distribution, DAVID considers genes with similar or related functions as a whole set for enrichment analysis.

## Results

**Figure 1:**
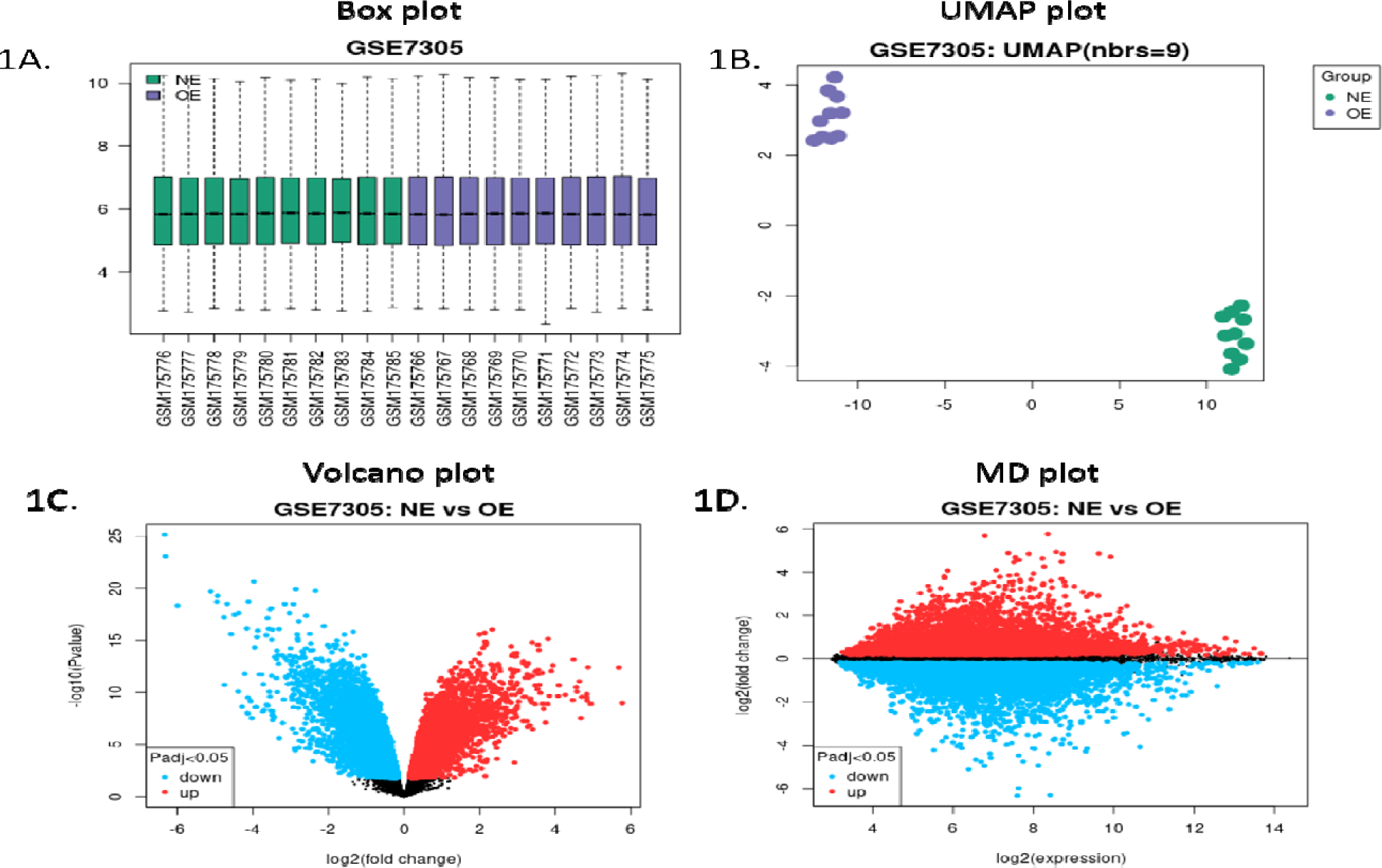
Data normalization and DEG’s visualization via Box plot (A), UMAP plot (B), Volcano plot (C) and MD plot (D).

**Figure 2:**
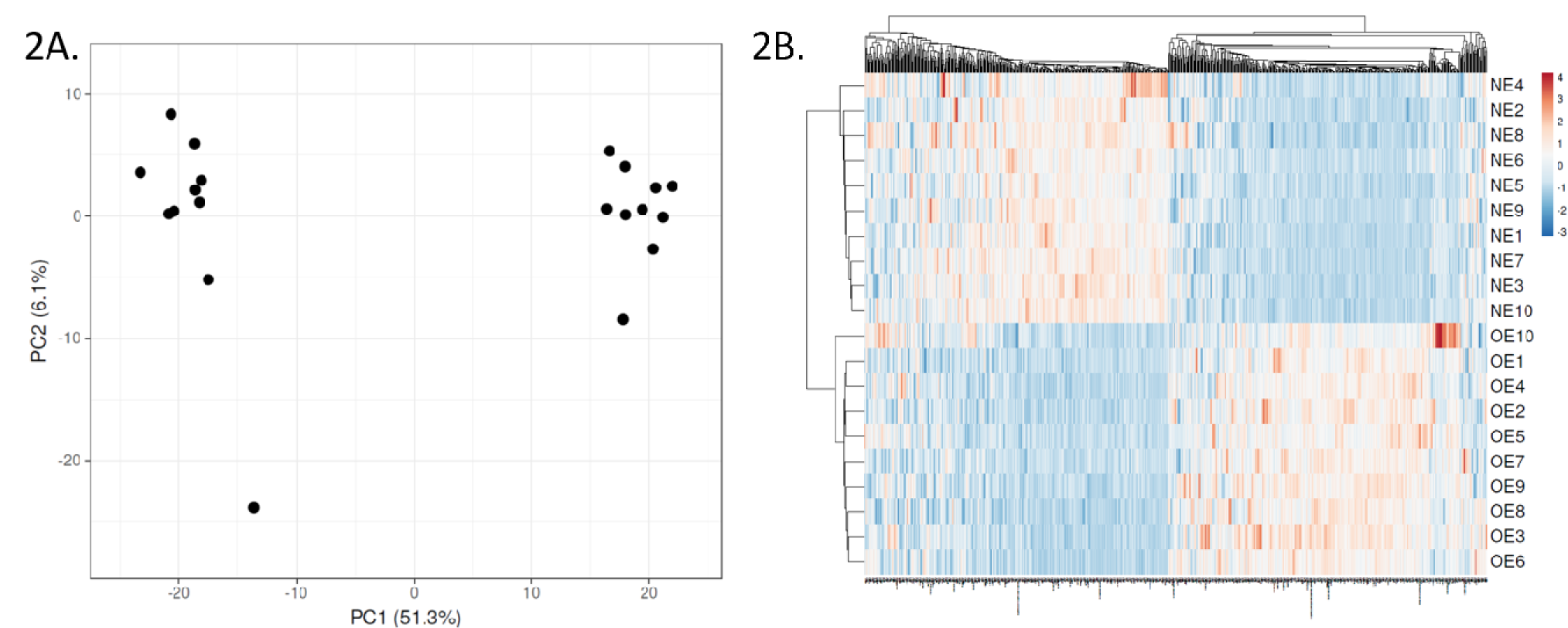
Heat map of DEG’s. Blue to orange gradation is for small to large changes in gene expression values.

**Figure 3:**
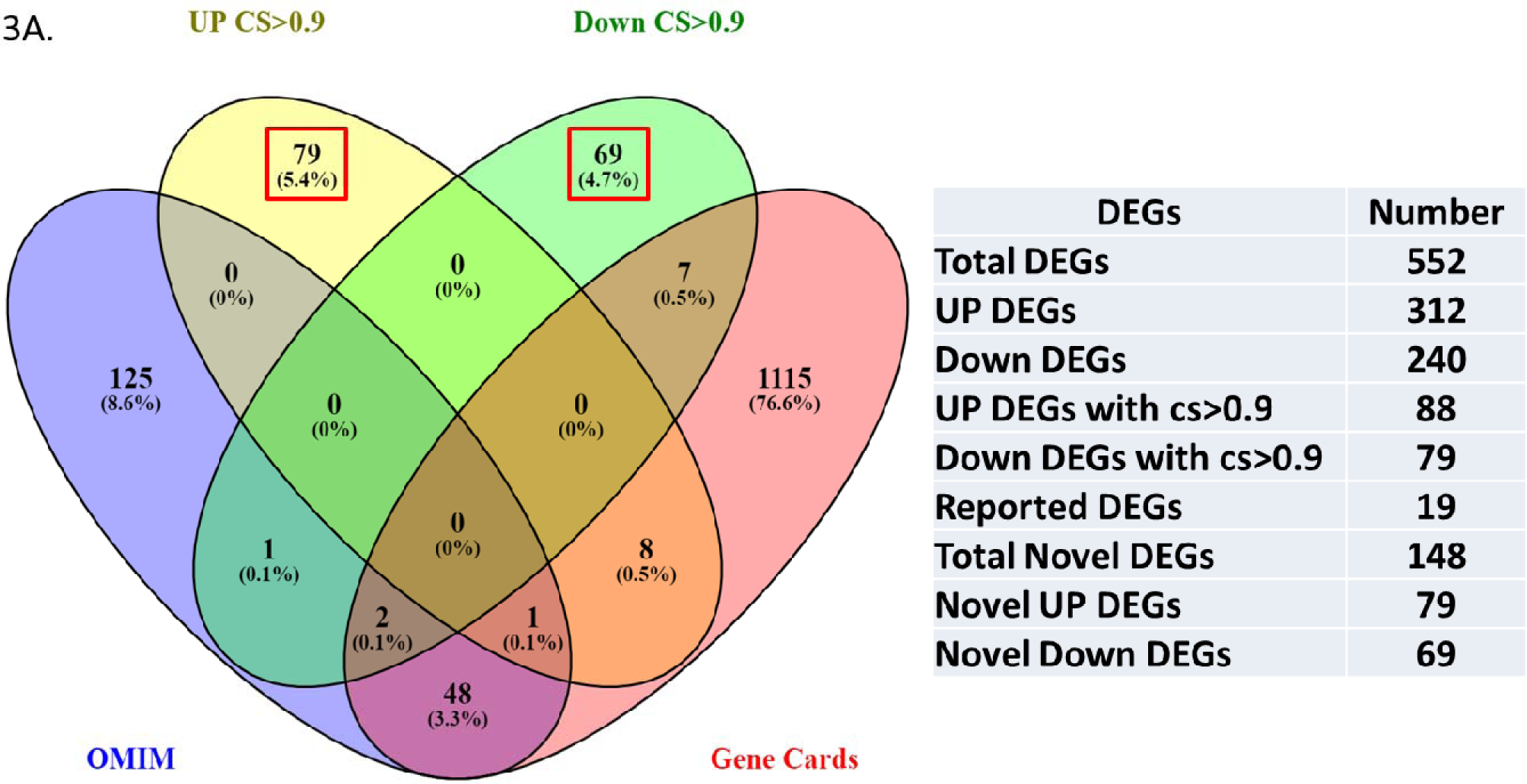
Venn diagram of the differentially expressed genes (DEGs).

### Differentially Expressed Genes (DEGs)

A total of 552 DEGs were identified with a p-value < 0.05, comprising 312 up-regulated and 240 down-regulated genes. Within this set, 148 DEGs were novel, consisting of 79 up-regulated and 69 down-regulated DEGs, selected based on their average gene expression values.

### Principal Component and Hierarchical Clustering Analysis of DEGs

Uniform Manifold Approximation and Projection (UMAP) plot exhibited distinct clustering of data with a neighborhood scoring of 9. The heat map, constructed for DEGs, visually represents the data matrix, where up-regulated DEGs are depicted in orange and down-regulated in blue. The color gradation from blue to orange signifies the numeric differences in gene expressions, ranging from small to large.

### Protein-Protein Interaction (PPI) Network

Based on the combined score calculated by STRING, a total of 297 gene pairs (combined score > 0.9) were found to interact, forming a primary network with 114 nodes and 237 edges (Figure 4). Additionally, four sub-networks were extracted. Hub nodes in the network included up-regulated DEGs such as C3, APP, GNG12, PSAP, GNAQ, and down-regulated DEGs like LPAR1, RAB3D, PIK3R1, TRIM32, CMTM6. Selection of hub nodes was based on clustering coefficient and edge betweenness criteria.

**Figure 4:**
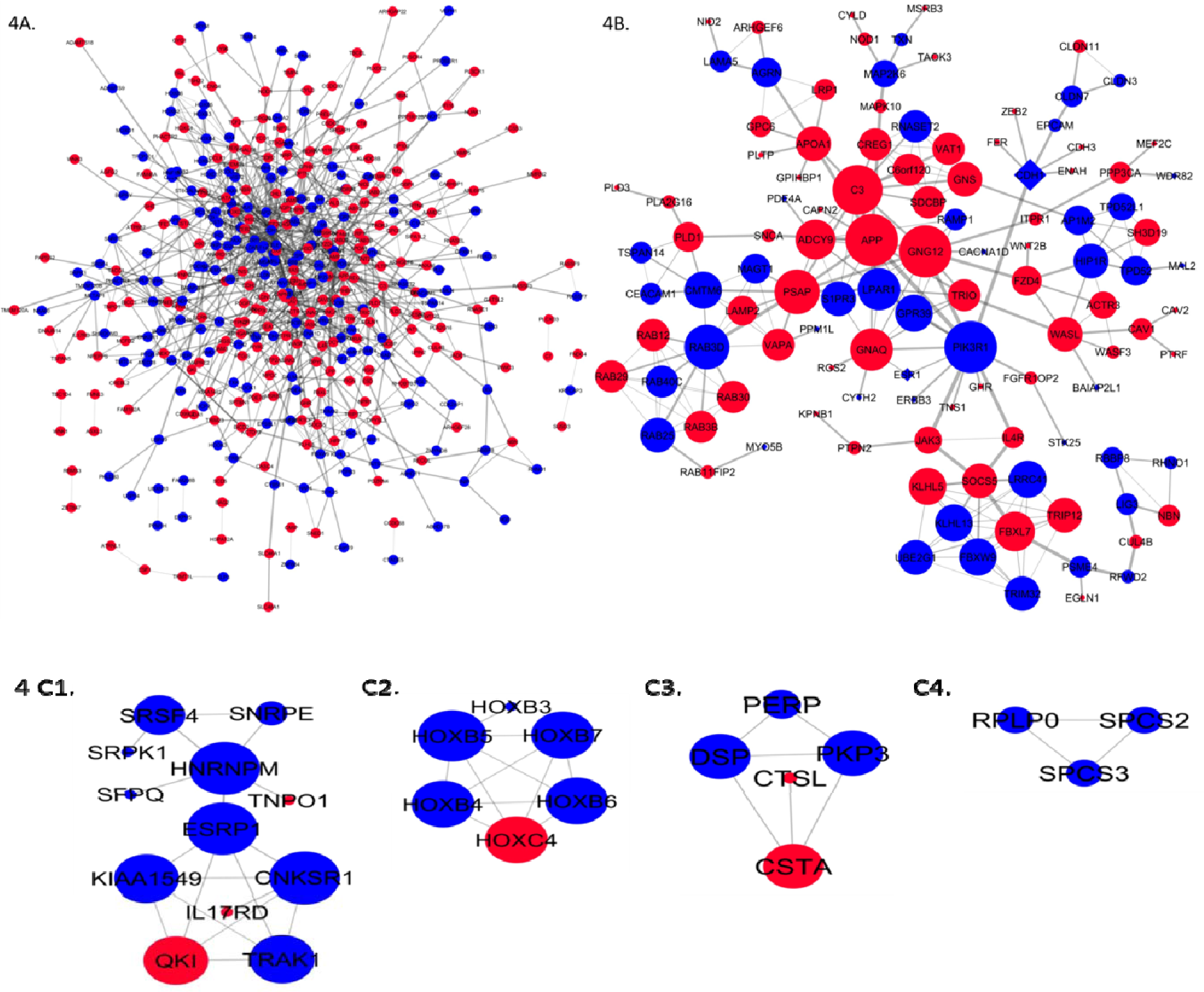
(A and B) Protein-protein interaction (PPI) of DEG’s. C1, C2, C3 and C4 are four sub-networks of DEG’s: C1; sub-network 1, C2; sub-network 2, C3; sub-network 3, C4; sub-network 4. Red-circle for up and blue-circle for down-regulated genes respectively. Blue diamond for similar related genes and lines shows the correlation between genes, where thickness of lines (edges), is proportional to the combined scores.

### Sub-network Extraction

Four sub-networks (C1, C2, C3, and C4) were extracted from the main network using Cytoscape (Fig. 4). In sub-network C4, all genes were down-regulated, whereas in sub-networks C1, C2, and C3, 31, 12, and 25 genes were up-regulated, respectively, and 9, 5, and 3 genes were down-regulated.

### Gene Ontology and Pathway Enrichment Analysis

Functional enrichment analysis was conducted, and major molecular functions, biological processes, and cellular components of DEGs with a false discovery rate (FDR) < 0.05 were listed in Tables 1 and 2. Results of GO enrichment analysis for upregulated DEGs (5A) identified hormone-mediated signaling, muscle fiber development, intracellular signaling cascade, and cellular protein complex assembly as major significant biological processes. Processes such as proteolysis, immune system development, epithelial cell differentiation, and development emerged as major significant biological processes related to downregulated DEGs (5B). Furthermore, KEGG pathway enrichment analysis (Fig. 5C) revealed ECM-receptor interaction and ubiquitin-mediated proteolysis as downregulated pathways. Upregulated pathways included melanogenesis, GnRH signaling pathway, and Alzheimer’s disease pathway.

**Figure 5:**
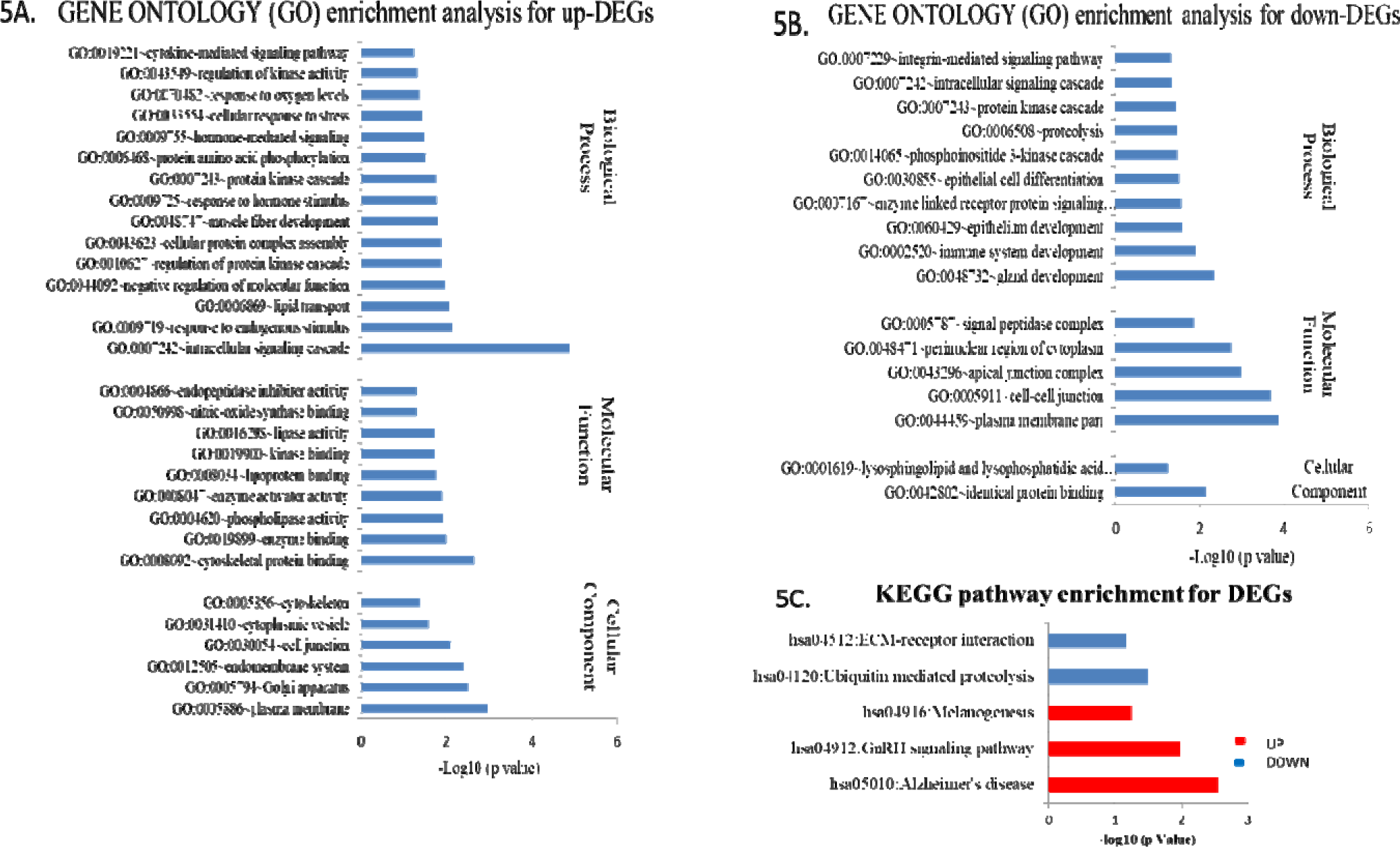
Gene Ontology (GO) enrichment analysis for up-regulated (5A) and down-regulated DEG’s (5B), and KEGG pathway enrichment for DEG’s (5C).

**Table 1:** GENE ONTOLOGY (GO) analysis for downregulated DEGs.

| Category | Term | P-Value |
| --- | --- | --- |
| GOTERM_BP_FAT | GO:0007242~intracellular signaling cascade | 1.34E-05 |
| GOTERM_BP_FAT | GO:0014706~striated muscle tissue development | 5.44E-04 |
| GOTERM_BP_FAT | GO:0042692~muscle cell differentiation | 5.87E-04 |
| GOTERM_BP_FAT | GO:0060537~muscle tissue development | 6.80E-04 |
| GOTERM_BP_FAT | GO:0042391~regulation of membrane potential | 9.31E-04 |
| GOTERM_BP_FAT | GO:0007517~muscle organ development | 0.001183 |
| GOTERM_BP_FAT | GO:0051146~striated muscle cell differentiation | 0.001501 |
| GOTERM_BP_FAT | GO:0010033~response to organic substance | 0.002202 |
| GOTERM_BP_FAT | GO:0048878~chemical homeostasis | 0.00222 |
| GOTERM_BP_FAT | GO:0010647~positive regulation of cell communication | 0.002453 |
| GOTERM_BP_FAT | GO:0006873~cellular ion homeostasis | 0.004966 |
| GOTERM_BP_FAT | GO:0006928~cell motion | 0.005073 |
| GOTERM_BP_FAT | GO:0055082~cellular chemical homeostasis | 0.00541 |
| GOTERM_BP_FAT | GO:0016044~membrane organization | 0.005487 |
| GOTERM_BP_FAT | GO:0007519~skeletal muscle tissue development | 0.00605 |
| GOTERM_BP_FAT | GO:0060538~skeletal muscle organ development | 0.00605 |
| GOTERM_BP_FAT | GO:0009967~positive regulation of signal transduction | 0.006229 |
| GOTERM_BP_FAT | GO:0009719~response to endogenous stimulus | 0.007589 |
| GOTERM_BP_FAT | GO:0050801~ion homeostasis | 0.007991 |
| GOTERM_BP_FAT | GO:0032273~positive regulation of protein polymerization | 0.008595 |
| GOTERM_BP_FAT | GO:0006869~lipid transport | 0.008949 |
| GOTERM_BP_FAT | GO:0042592~homeostatic process | 0.009037 |
| GOTERM_BP_FAT | GO:0048741~skeletal muscle fiber development | 0.010712 |
| GOTERM_BP_FAT | GO:0044092~negative regulation of molecular function | 0.011126 |
| GOTERM_BP_FAT | GO:0001765~membrane raft formation | 0.011205 |
| GOTERM_BP_FAT | GO:0045807~positive regulation of endocytosis | 0.011464 |
| GOTERM_BP_FAT | GO:0010876~lipid localization | 0.011736 |
| GOTERM_BP_FAT | GO:0052547~regulation of peptidase activity | 0.012493 |
| GOTERM_BP_FAT | GO:0010627~regulation of protein kinase cascade | 0.012994 |
| GOTERM_MF_FAT | GO:0008092~cytoskeletal protein binding | 0.002399 |
| GOTERM_MF_FAT | GO:0016504~peptidase activator activity | 0.007702 |
| GOTERM_MF_FAT | GO:0019899~enzyme binding | 0.010389 |
| GOTERM_MF_FAT | GO:0003779~actin binding | 0.01129 |
| GOTERM_MF_FAT | GO:0004620~phospholipase activity | 0.012226 |
| GOTERM_MF_FAT | GO:0008047~enzyme activator activity | 0.012783 |
| GOTERM_MF_FAT | GO:0008289~lipid binding | 0.015001 |
| GOTERM_MF_FAT | GO:0008034~lipoprotein binding | 0.017323 |
| GOTERM_MF_FAT | GO:0019900~kinase binding | 0.019893 |
| GOTERM_MF_FAT | GO:0016298~lipase activity | 0.020073 |
| GOTERM_MF_FAT | GO:0070290~NAPE-specific phospholipase D activity | 0.028556 |
| GOTERM_MF_FAT | GO:0042802~identical protein binding | 0.030878 |
| GOTERM_MF_FAT | GO:0070064~proline-rich region binding | 0.03417 |
| GOTERM_MF_FAT | GO:0008035~high-density lipoprotein binding | 0.039752 |
| GOTERM_MF_FAT | GO:0004630~phospholipase D activity | 0.039752 |
| GOTERM_MF_FAT | GO:0008139~nuclear localization sequence binding | 0.039752 |
| GOTERM_MF_FAT | GO:0019904~protein domain specific binding | 0.042369 |
| GOTERM_MF_FAT | GO:0050998~nitric-oxide synthase binding | 0.050821 |
| GOTERM_MF_FAT | GO:0004866~endopeptidase inhibitor activity | 0.05148 |
| GOTERM_MF_FAT | GO:0030414~peptidase inhibitor activity | 0.058629 |
| GOTERM_CC_FAT | GO:0030055~cell-substrate junction | 4.14E-04 |
| GOTERM_CC_FAT | GO:0005886~plasma membrane | 0.001132 |
| GOTERM_CC_FAT | GO:0005912~adherens junction | 0.001789 |
| GOTERM_CC_FAT | GO:0005626~insoluble fraction | 0.002358 |
| GOTERM_CC_FAT | GO:0005925~focal adhesion | 0.002595 |
| GOTERM_CC_FAT | GO:0070161~anchoring junction | 0.002815 |
| GOTERM_CC_FAT | GO:0005924~cell-substrate adherens junction | 0.002983 |
| GOTERM_CC_FAT | GO:0005794~Golgi apparatus | 0.00324 |
| GOTERM_CC_FAT | GO:0009986~cell surface | 0.003371 |
| GOTERM_CC_FAT | GO:0012505~endomembrane system | 0.004123 |
| GOTERM_CC_FAT | GO:0031982~vesicle | 0.004141 |
| GOTERM_CC_FAT | GO:0005624~membrane fraction | 0.005317 |
| GOTERM_CC_FAT | GO:0048471~perinuclear region of cytoplasm | 0.00559 |
| GOTERM_CC_FAT | GO:0016323~basolateral plasma membrane | 0.00569 |
| GOTERM_CC_FAT | GO:0044431~Golgi apparatus part | 0.006169 |
| GOTERM_CC_FAT | GO:0030027~lamellipodium | 0.007163 |
| GOTERM_CC_FAT | GO:0044459~plasma membrane part | 0.008355 |
| GOTERM_CC_FAT | GO:0030054~cell junction | 0.008464 |
| GOTERM_CC_FAT | GO:0009925~basal plasma membrane | 0.009319 |
| GOTERM_CC_FAT | GO:0045178~basal part of cell | 0.012293 |
| GOTERM_CC_FAT | GO:0031088~platelet dense granule membrane | 0.016805 |
| GOTERM_CC_FAT | GO:0000267~cell fraction | 0.017154 |
| GOTERM_CC_FAT | GO:0044421~extracellular region part | 0.017907 |
| GOTERM_CC_FAT | GO:0000139~Golgi membrane | 0.020736 |
| GOTERM_CC_FAT | GO:0031410~cytoplasmic vesicle | 0.027418 |
| GOTERM_CC_FAT | GO:0005955~calcineurin complex | 0.027853 |
| GOTERM_CC_FAT | GO:0042827~platelet dense granule | 0.033332 |
| GOTERM_CC_FAT | GO:0005901~caveola | 0.03462 |
| GOTERM_CC_FAT | GO:0016023~cytoplasmic membrane-bounded vesicle | 0.034768 |
| GOTERM_CC_FAT | GO:0005615~extracellular space | 0.038156 |
| GOTERM_CC_FAT | GO:0031988~membrane-bounded vesicle | 0.040294 |
| GOTERM_CC_FAT | GO:0042995~cell projection | 0.0416 |
| GOTERM_CC_FAT | GO:0031252~cell leading edge | 0.042819 |
| GOTERM_CC_FAT | GO:0005856~cytoskeleton | 0.043217 |
| GOTERM_CC_FAT | GO:0005576~extracellular region | 0.051766 |
| GOTERM_CC_FAT | GO:0019898~extrinsic to membrane | 0.059319 |

**Table 2:**
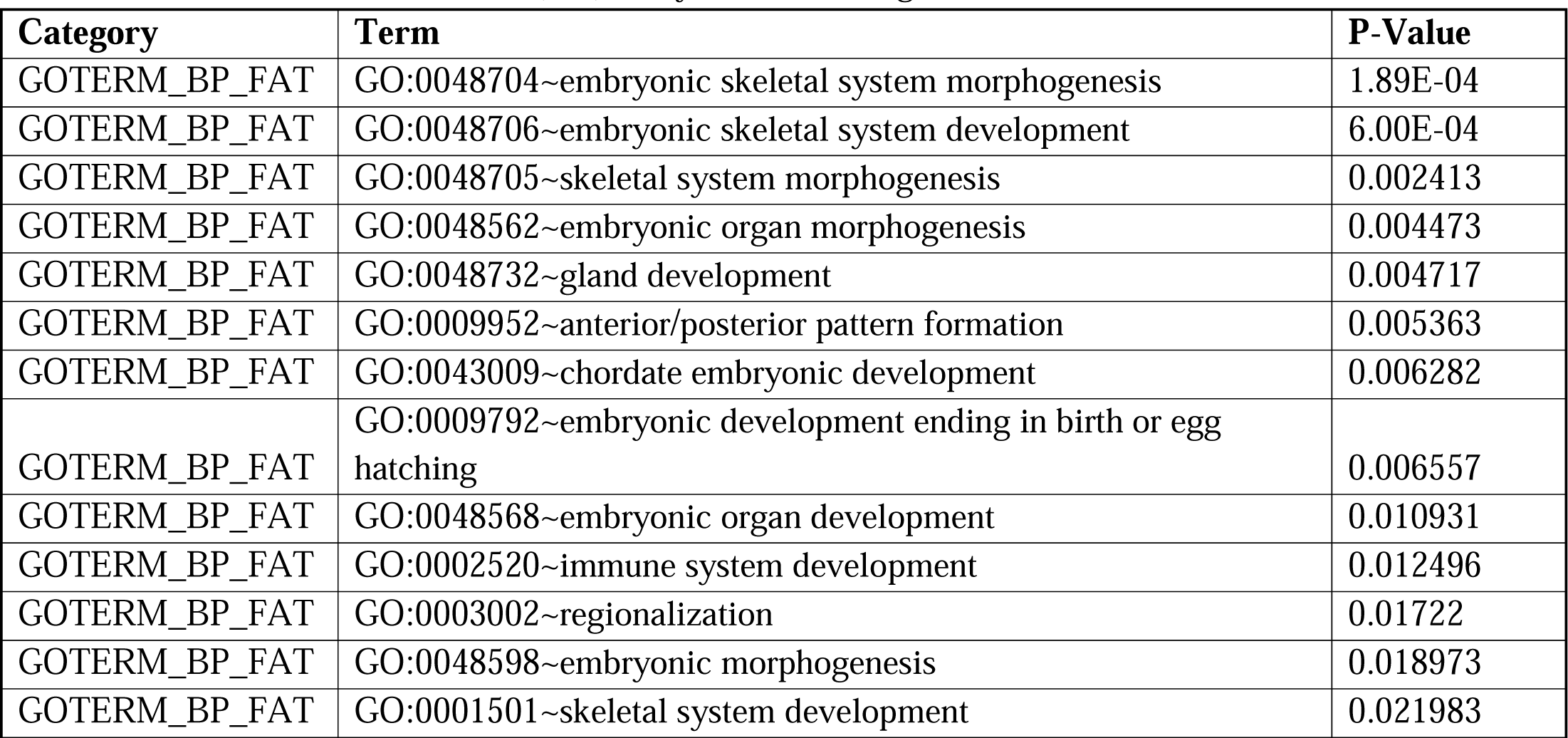
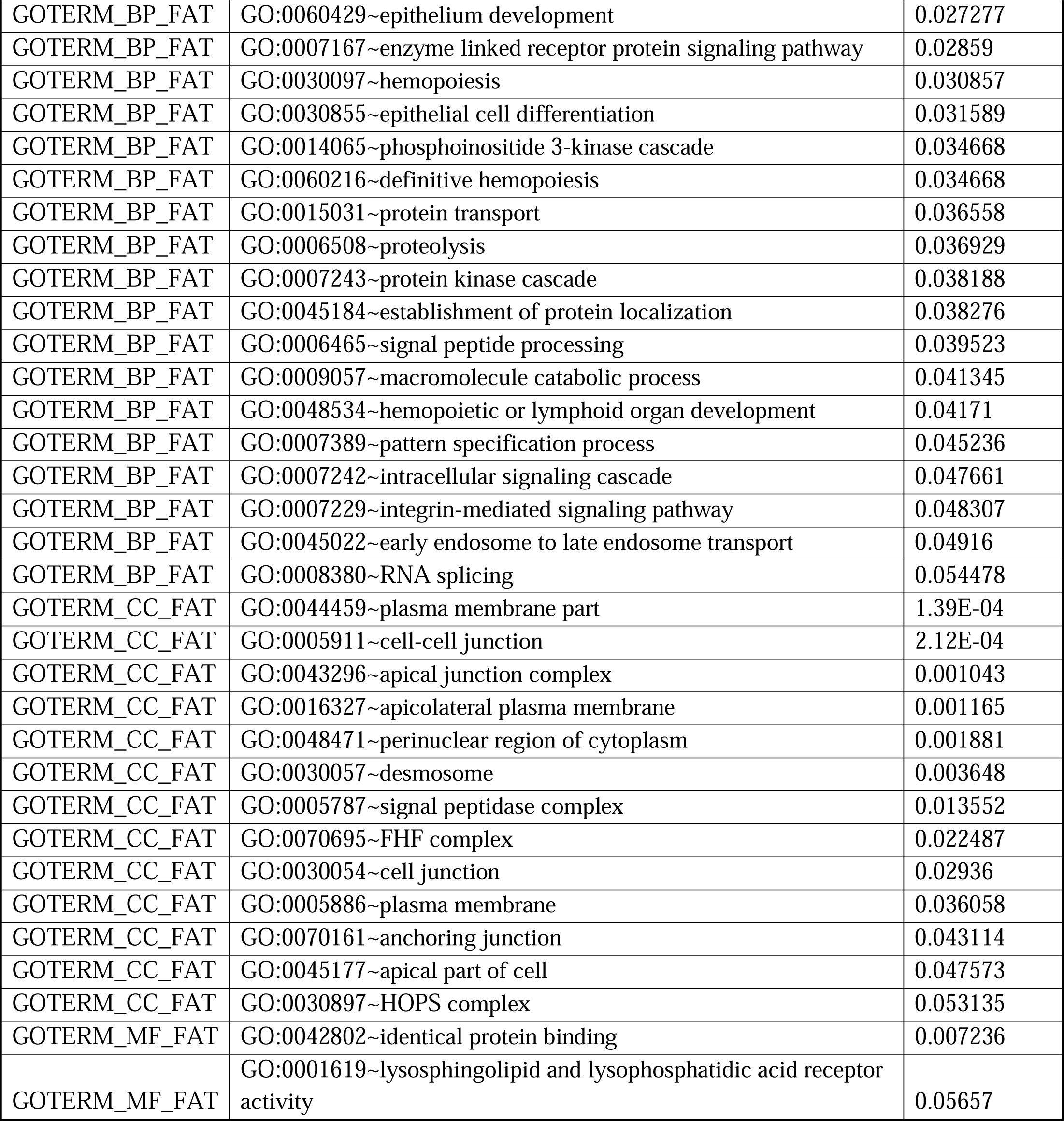
GENE ONTOLOGY (GO) analysis for down regulated DEGs.

| Category | Term | P-Value |
| --- | --- | --- |
| GOTERM_BP_FAT | GO:0048704~embryonic skeletal system morphogenesis | 1.89E-04 |
| GOTERM_BP_FAT | GO:0048706~embryonic skeletal system development | 6.00E-04 |
| GOTERM_BP_FAT | GO:0048705~skeletal system morphogenesis | 0.002413 |
| GOTERM_BP_FAT | GO:0048562~embryonic organ morphogenesis | 0.004473 |
| GOTERM_BP_FAT | GO:0048732~gland development | 0.004717 |
| GOTERM_BP_FAT | GO:0009952~anterior/posterior pattern formation | 0.005363 |
| GOTERM_BP_FAT | GO:0043009~chordate embryonic development | 0.006282 |
| GOTERM_BP_FAT | GO:0009792~embryonic development ending in birth or egg hatching | 0.006557 |
| GOTERM_BP_FAT | GO:0048568~embryonic organ development | 0.010931 |
| GOTERM_BP_FAT | GO:0002520~immune system development | 0.012496 |
| GOTERM_BP_FAT | GO:0003002~regionalization | 0.01722 |
| GOTERM_BP_FAT | GO:0048598~embryonic morphogenesis | 0.018973 |
| GOTERM_BP_FAT | GO:0001501~skeletal system development | 0.021983 |
| GOTERM_BP_FAT | GO:0060429~epithelium development | 0.027277 |
| GOTERM_BP_FAT | GO:0007167~enzyme linked receptor protein signaling pathway | 0.02859 |
| GOTERM_BP_FAT | GO:0030097~hemopoiesis | 0.030857 |
| GOTERM_BP_FAT | GO:0030855~epithelial cell differentiation | 0.031589 |
| GOTERM_BP_FAT | GO:0014065~phosphoinositide 3-kinase cascade | 0.034668 |
| GOTERM_BP_FAT | GO:0060216~definitive hemopoiesis | 0.034668 |
| GOTERM_BP_FAT | GO:0015031~protein transport | 0.036558 |
| GOTERM_BP_FAT | GO:0006508~proteolysis | 0.036929 |
| GOTERM_BP_FAT | GO:0007243~protein kinase cascade | 0.038188 |
| GOTERM_BP_FAT | GO:0045184~establishment of protein localization | 0.038276 |
| GOTERM_BP_FAT | GO:0006465~signal peptide processing | 0.039523 |
| GOTERM_BP_FAT | GO:0009057~macromolecule catabolic process | 0.041345 |
| GOTERM_BP_FAT | GO:0048534~hemopoietic or lymphoid organ development | 0.04171 |
| GOTERM_BP_FAT | GO:0007389~pattern specification process | 0.045236 |
| GOTERM_BP_FAT | GO:0007242~intracellular signaling cascade | 0.047661 |
| GOTERM_BP_FAT | GO:0007229~integrin-mediated signaling pathway | 0.048307 |
| GOTERM_BP_FAT | GO:0045022~early endosome to late endosome transport | 0.04916 |
| GOTERM_BP_FAT | GO:0008380~RNA splicing | 0.054478 |
| GOTERM_CC_FAT | GO:0044459~plasma membrane part | 1.39E-04 |
| GOTERM_CC_FAT | GO:0005911~cell-cell junction | 2.12E-04 |
| GOTERM_CC_FAT | GO:0043296~apical junction complex | 0.001043 |
| GOTERM_CC_FAT | GO:0016327~apicolateral plasma membrane | 0.001165 |
| GOTERM_CC_FAT | GO:0048471~perinuclear region of cytoplasm | 0.001881 |
| GOTERM_CC_FAT | GO:0030057~desmosome | 0.003648 |
| GOTERM_CC_FAT | GO:0005787~signal peptidase complex | 0.013552 |
| GOTERM_CC_FAT | GO:0070695~FHF complex | 0.022487 |
| GOTERM_CC_FAT | GO:0030054~cell junction | 0.02936 |
| GOTERM_CC_FAT | GO:0005886~plasma membrane | 0.036058 |
| GOTERM_CC_FAT | GO:0070161~anchoring junction | 0.043114 |
| GOTERM_CC_FAT | GO:0045177~apical part of cell | 0.047573 |
| GOTERM_CC_FAT | GO:0030897~HOPS complex | 0.053135 |
| GOTERM_MF_FAT | GO:0042802~identical protein binding | 0.007236 |
| GOTERM_MF_FAT | GO:0001619~lysosphingolipid and lysophosphatidic acid receptor activity | 0.05657 |

## Discussion

Endometriosis, a complex gynecological disease, significantly impacts the reproductive health of women. Our study aimed to unravel the molecular mechanisms underlying the progression of endometriosis, utilizing comprehensive analysis of gene expression datasets through various bioinformatics tools.

The examination of gene expression profiles led to the identification of 552 differentially expressed genes (DEGs), including 312 up-regulated and 240 down-regulated genes. The up-regulated DEGs were found to be associated with essential biological processes such as cytokine-mediated signaling pathway, cellular response to stress, hormone-mediated signaling, lipid transport, and response to endogenous stimuli. Molecular functions of these up-regulated DEGs included lipase, kinase, lipoprotein, enzyme and cytoskeleton protein binding, enzyme activator, and phospholipase activity. They were also integral components of cellular structures such as the cytoskeleton, cytoplasmic vesicle, cell junctions, endomembrane system, Golgi apparatus, and plasma membrane.

Conversely, down-regulated DEGs were implicated in biological processes like proteolysis, gland and immune system development, integrin-mediated signaling, intracellular signaling, and protein kinase cascade. Their molecular functions involved signal peptidase, apical junction, cell-cell junction complex, perinuclear regions, and plasma membrane, with cellular components related to lysosphingolipid, lysophosphatidic acid, and identical protein binding.

Key hub nodes in the protein-protein interaction network were identified, with up-regulated DEGs including C3, APP, GNG12, PSAP, and GNAQ, while LPAR1, RAB3D, PIK3R1, TRIM32, and CMTM6 represented down-regulated DEGs. These hub nodes play crucial roles in the intricate molecular landscape of endometriosis.

**C3** (Complement Component 3) is one of the important complement proteins out of 30 recognized till date. C3 has intensive and pivotal role in complement activation in both classical and alternative pathways. The alternative pathway is independent of antigen-antibody complexes and can directly be induced by components of cell wall of bacteria or present on the surface of damaged host cell via C3 unlike classical and lectin pathway (19). Altered immune system is among the various risk factors which are involved in pathophysiology of endometriosis and hence deregulated C3 (which is an important player of immune system) might be involved in the progression of endometriosis and can be considered as a potent biomarker.

**APP** (Amyloid precursor protein), plays an important role in synaptic activity and neuronal plasticity, but upto this time it’s not completely revealed (20). APP gene has been found to be associated with down’s syndrome (21). Mutation in APP gene were also found to be associated with dementia and Alzimers disease including amyloid deposition, neurofibrillary tangle formation and cerebral amyloid angiopathy (CAA)(22). APP is involved in the proliferation, migration and adhesion of endothelial cells. It mediates stability to focal adhesion and cell-cell junctions, while its necessary for VEGF-A growth factor responses (23).Thus, it can be said that APP may be a factor to be involved in the adhesion and cellular junction formation of endometrial tissues during endometriosis.

**GNG12** is known as the c12 subunit of G protein, and G proteins are made by three different subunits a, b and c making heterotrimeric. The different subunits are interchangeable, making possible combinations and wide array of effects. GNG12 is highly conserved, as and its homologs are present in human, rat, cow, frog, chicken and zebrafish. C12 is expressed differentially in mammalian brain, where its localized in glial cells and expressed in reactive astrocytes (24).

**PSAP (Prosaposin,** also known as SGP-1) is an intriguing multifunctional protein that plays roles intracellularly in regulation of lysosomal enzyme functions, and extracellularly, as a secretary factor having both neuroprotective and glioprotective effects. PSAP gene encodes a 524 amino-acid precursor protein prosaposin (pSAP), that gives 4 small glycoproteins-saposins (SapA, B, C and D) (25). Saposins, a sphingolipid activator protein that’s required for the function of lysosomal hydrolases. Mutation in PSAP gene (two null alleles) in an individual have pSap deficiency and suffer with fatal infantile lysosomal storage disease(26). Defects of Sap A and SapC leads to atypical Krabbe and Gaucher disease (27). Any defect in SapB results in MLD (metachromatic leukodystrophy) due to impaired degradation and accumulation of cerebroside-3 sulfate (sulfatide) (28), while SapD deficiency causes Farber disease in mice (29).

**GNAQ (**Gnaq) is a member of guanine nucleotide binding protein (G protein) subunits and is found abundantly in brain. Gnaq gene knocking shows serious nervous dysfunction and endocrine system in mice (30) .Gnaq gene mutation has been reported mostly in uveal melanoma (31). Gnaq, Gq protein a-subunit, encoded by GNAQ gene, is a member of Gq/11subfamily of heterotrimeric G proteins and its ubiquitously expressed in mammalian cells (30,32). GNAQ has role in cardiovascular system (31), cancer and autoimmune diseases (33). It is found to be coupled with GPCRs in viral infections(34), but no any expression or mutation studies has been yet reported in endometriosis, adenomyosis or ovarian carcinoma.

### PIK3R1

PI3K enzymes are lipid kinases, having a conserved sequences and phosphorylating the inositol 30-OH groups of membrane phosphoinositides (PI).

Class I PI3K convert phosphoinositol bisphosphate (PIP2) (4,5) into phosphoinositol trisphosphate (PIP3) (3,4,5) a second messenger (35). Class IA PI3K is composed of a heterodimer having a p110 catalytic subunit and a p85 regulatory subunit. Out of four PI3K catalytic subunit isoforms (PI3Ka, PI3Kb, PI3Kg, and PI3Kd), only PI3Ka and PI3Kb are ubiquitously expressed in the body and are frequently altered in cancer disease (36). Three different genes PIK3R1, PIK3R2, and PIK3R3, encode p85-type subunits; p85a, p85b and p55g, respectively. The two of PIK3R1 and PIK3R2 are widely expressed in the body, whereas the third one PIK3R3 is expressed only in testis and brain of adults (37). PIK3R1/p85a is isoform found abundantly, but in cancer patients its expression is reduced (38). It’s a Tumor-Suppressor Gene and the most striking difference between p85a and p85b is that PIK3R1/p85a acts as a tumor suppressor while, PIK3R2/p85b is a cancer driver. The recent finding say that p85a subunit restrains the catalytic activity of PI3K (39) encourage testing the consequences of reducing p85a levels. Other study tells deletion of Pik3r1 led to a gradual change in hepatocyte morphology (liver) and with over time, mice develops hepatocellular carcinoma (40). These two observations confirm the deletion of Pik3r1 gene is a cause of tumor development, as similarly observed for tumor suppressors. Pik3r1 loss in mouse accelerates HER2/neu-induced mammary cancer development, in cultured human epithelial cells PIK3R1 knockdown cause transformational changes, while hemi-zygous deletion is a frequent event in breast cancer samples (41). Mutation in PIK3R1 has been reported in breast, pancreatic, colon cancers (42)and in 8.5% cases of ovarian carcinoma (43) The PIK3 pathway is downstream regulated from RTK (receptor tyrosine kinase), that’s active in cancer lineages, including endometrial cancer (EC) (44). PIK3 mutation has been reported in ovarian carcinoma but not in cases where patients suffer with endometriosis only(45). While PI3K pathway has been found to be strongly implicated during the development of endometriosis (46).

### LPAR1

Lysophosphatidic Acid, is a small phospholipid present in many mammalian cells and tissues (47). It is involved in cell migration, survival, proliferation, cellular interactions and cytoskeleton changes (48). A study on mare, suffering with endometriosis has indicated that concentration of LPA, its receptors, PGE2/PGF2 ration and CTGF secretion is altered during endometriosis (49). LPA induces IL-8 (Interleukin-8) expression via LPA-1 receptor, Gi protein, MAPK/p38 and NF-kB signalling pathway. Where IL-8 protein stimulates endometrial cell migration, permeability, capillary tube formation and proliferation leading to angiogenesis during pregnancy (50). Overexpression of all LPARs and enzymes occur responsible for LPA synthesis, during endometrial cancers, showing a positive correlation with myoinvasion and FIGO (International Federation of Gynecology and Obstetrics) stage (51). Recent study has demonstrated that LPA, acts as a mitogen and pro-invasive stimulus for endometrial and endometriotic cells acts via LPAR1 and LPAR3 (Lysophosphatidic Acid Receptors). It has demonstrated that LPA-dependent stimulus causes secretion of cathepsin B, a protease, which acts as a factor for endometriotic invasion (52).

**RAB3D,** is a GTPase of Rab family, plays a role as a central regulator of vesicular transport (53).A study reveals that the Rab3D is implicated in the subcellular localization and maturation of Immature Secretory granules (ISGs) (54). Rabs are involved in membrane trafficking, cellular signalling, growth and development. Rabs and their effectors has been found to be overexpressed or undergone for loss of function in many disease including cancers and causes the disease progression(55)

.Endometriosis is also an inflammatory disease and similarly Rab can play its role in endometriosis disease progression and adhesion.

**GPR39,** are G-protein coupled receptors present in the plasma membrane, which is distinct target for binding of extracellular Zn^+^ ions (56–58). G-protein coupled receptors are a large family of seven-transmembrane proteins, which are all involved in a diverse array of extracellular stimuli (59). Zinc is an important component of enzymes and proteins. It is also required for intracellular message transmission, protein synthesis, cell membrane maintenance and transport, regulation of neuronal, endocrinal system (60). In brain GPR39 zinc sensing receptor has a significant role in Alzheimer and Epilepsy (61). Further investigations showed that GPR39 contributes its part in skin wound healing, thus having a positive role in the area of dermatology and stem cell therapy (62). A study has revealed that GPR39 might be a direct promising target for therapy of zincergic dyshomeostasis observed in Alzheimer’s disease, but more studies are still needed over it (63).

**HOXC4**, is actively transcribed during the development and differentiation of Lymphoid, myeloid and erythroid cells. It helps to maintain proliferation in hematopoietic cells(64).

Cystatin A (**CSTA**), is a type 1 cystatin super-family member, and is expressed mainly in epithelial and lymphoid tissues. It prevents the proteolysis of cytoplasmic and cytoskeletal proteins in cells. It acts a tumor suppressor in esophageal and lung cancer (65). The risk of disease recurrence and death are found to be higher in patients suffering with squamous cell carcinoma of head and neck, having low CSTA(66) .Here expression analysis of DEGs in endometriosis, CSTA was found to be downregulated but detailed studies for the role of CSTA in endometriosis disease progression is still has to be done.

Epithelial splicing regulatory protein 1 (**ESRP1**), plays crucial role during organogenesis of craniofacial and epidermal development, branching morphogenesis in the lungs and salivary glands. It also plays role during cancer progression, and its expression found to be low in normal epithelium but upregulated during carcinoma (67).

## Conclusion

This study has unveiled a diverse array of genes that likely play pivotal roles, directly or indirectly, in the pathophysiology of endometriosis. The identified genes contribute to the intricate molecular landscape involved in the progression of the disease. It is evident that multiple genes collaboratively participate in shaping the trajectory of endometriosis.While this study provides valuable insights into the physiological aspects of the disease, it is important to acknowledge the complexity of endometriosis and the likelihood of numerous genes collectively influencing its progression. The findings serve as a stepping stone towards a better understanding of the disease, yet a more detailed and comprehensive study is imperative to unravel the nuanced mechanisms underlying endometriosis fully.This study lays the groundwork for further exploration, encouraging future investigations to delve deeper into the roles and interactions of these genes. Such endeavors will contribute to the development of targeted therapeutic interventions and more accurate diagnostic strategies for endometriosis.

## Ethics statement

The studies involving human participants were reviewed and approved by Ethics Committee of the Institutional Ethical committee before starting the study (No. Dean/2018/EC/936).

## Author contributions

RS, RCand SR conceived and designed the project. KK and AA performed all operations. RB analyzed the data and drew the figures. KK and RB wrote the manuscript. AA, KK, RB and RS revised the manuscript. All authors contributed to the article and approved the submitted version.

## Data Availability

All data produced in the present work are contained in the manuscript

## Acknowledgement

We want to extend our sincere gratitude to Multi-Disciplinary Research Units (MRUs) Laboratory, a grant by ICMR-Department of Health Research.

